# A Natural Language Processing-Based Approach for Early Detection of Heart Failure Onset using Electronic Health Records

**DOI:** 10.1101/2025.04.04.25325211

**Authors:** Yuxi Liu, Zhen Tan, Zhenhao Zhang, Song Wang, Jingchuan Guo, Huan Liu, Tianlong Chen, Jiang Bian

## Abstract

**Objectives:** This study set out to develop and validate a risk prediction tool for the early detection of heart failure (HF) onset using real-world electronic health records (EHRs).

**Background:** While existing HF risk assessment models have shown promise in clinical settings, they are often tailored to specific medical conditions, limiting their generalizability. Moreover, most methods rely on hand-crafted features, making it difficult to capture the high-dimensional, sparse, and temporal nature of EHR data, thus reducing their predictive accuracy.

**Methods:** A total of 2,561 HF and 5,493 matched control patients were identified from the OneFlorida Clinical Research Consortium. We employed a suite of natural language processing (NLP) models, including Bag of Words, Skip-gram, and ClinicalBERT, to generate EHR embeddings, which were used as inputs for five prediction models. Model calibration was assessed under three calibration scenarios: no recalibration, recalibration in the large, and logistic recalibration.

**Results:** The XGBoost model demonstrated the best overall performance, achieving an AUROC of 0.7672, an F1 score of 0.5547, an AUPRC of 0.6382, and a Matthews correlation coefficient of 0.3993. The most impactful predictors included diagnoses, procedures, medications, lab tests, and patient age. Model performance varied across gender, race, and ethnicity subgroups. Logistic recalibration significantly improved model calibration in the overall cohort and demographic subgroups.

**Conclusions:** Our NLP-based approach demonstrated strong predictive performance and clinical relevance, highlighting its potential for integration into real-world clinical applications to facilitate early detection and proactive management of individuals at risk for HF.

## 1. Introduction

Heart failure (HF) is an alarmingly increasing public health concern, affecting an estimated 26 million individuals worldwide [1]. As a chronic and progressive syndrome, HF is characterized by a reduced capacity of the heart to pump and fill with blood, resulting in inadequate circulation that fails to meet the metabolic demands of the body [2].

In the United States, the prevalence of HF is increasing at an alarming rate. According to data from the 2017-2020 National Health and Nutrition Examination Survey, an estimated 6.7 million adults aged 20 years and older are currently living with HF [3], with predictions suggesting an increase to 8.5 million by 2030 [4]. The economic burden associated with HF is also substantial and is expected to grow significantly over time. For instance, total direct medical costs in the United States are expected to increase from $21 billion in 2012 to $53 billion by 2030, while total costs, including indirect costs, may increase from $31 billion to $70 billion [3]. Besides, HF is associated with high mortality, especially within the first year of diagnosis in older adults [5–7]. Therefore, there is an urgent need to develop targeted prevention strategies to mitigate the growing burden of HF on public health and healthcare systems.

Predictive analytics in healthcare has emerged as a promising strategy to address this need by enabling early identification of at-risk individuals, reducing treatment costs, and improving patient outcomes [8]. Traditionally, machine learning and statistical models using electronic health records (EHRs) have been used to estimate disease outcomes and assess patient risk profiles [9–12]. Recently, the advent of deep learning techniques, particularly transformer-based architectures [13], has significantly advanced the field. Transformer is a type of neural network specifically designed to handle sequential data through self-attention mechanisms that capture long-range dependencies and contextual relationships within the input. Bidirectional encoder representations from transformers (BERT) is currently the most popular method for dealing with EHR data [14]. The BERT method facilitates the integration and modeling of high-dimensional sparse EHR data, capturing complex patterns and temporal dynamics in patient data [15–18]. A key factor contributing to this progress is transfer learning, which was originally introduced in the field of natural language processing (NLP) and allows models to be pretrained on large-scale unannotated datasets and fine-tuned on specific downstream tasks. These capabilities establish transformer-based models as scalable solutions for secondary healthcare applications, capable of evolving alongside new data and clinical contexts. Accordingly, the integration of deep learning into predictive analytics offers new opportunities for enhancing clinical decision support systems and promoting personalized and proactive care strategies.

In this study, we conducted a comprehensive predictive analysis aimed at the early detection of HF onset using EHR data collected from a single medical institution. We employed a suite of NLP models to generate dense vector representations for structured medical codes. Specifically, we explored three types of models: (1) Bag-of-Words (BoW), which encodes the frequency of code occurrences; (2) Skip-gram, which captures local co-occurrence patterns by predicting surrounding codes within a fixed context window; and (3) ClinicalBERT [16], a transformer-based language model pre-trained on large-scale clinical corpora, capable of generating contextualized embeddings.

Unlike static embeddings, ClinicalBERT captures semantic relationships by leveraging the sequential and contextual dependencies inherent in patient data. These embeddings are fed into machine learning models to assess their effectiveness in identifying patients at high risk of developing HF. To further enhance the clinical utility of our approach, we conducted a detailed analysis of feature importance to identify the key risk factors affecting model decisions. We evaluated model performance across gender, race, and ethnicity subgroups to account for potential disparities. We carried out a rigorous calibration analysis to examine the alignment between predicted risks and observed outcomes.

## 2. Methods

### 2.1. Data description

This study utilized data from the OneFlorida Clinical Research Consortium, part of the national PCORnet initiative. The dataset comprises EHRs merged with various sources, including Medicaid and Medicare claims, vital statistics, and selected tumor registries, covering approximately 16.8 million residents of Florida since 2012. It provides comprehensive patient information such as demographics, diagnoses, medications, procedures, vital signs, and lab tests. The study was approved by the University of Florida Institutional Review Board under IRB202201080.

### 2.2. Candidate variables

In this study, we developed predictive models aimed at the early detection of HF onset by using a comprehensive set of candidate variables derived from EHR data. Demographic data from patients, including age, sex, race, and ethnicity, were included due to their established associations with the risk and progression of HF [19]. We incorporated diagnosis codes (i.e., International Classification of Diseases, Tenth Revision (ICD-10)), procedure codes (i.e., Current Procedural Terminology (CPT)), medications (i.e., the 11-digit National Drug Code (NDC) and RxNorm Concept Unique Identifiers (RXCUIs)), vital signs (i.e., diastolic and systolic blood pressure (DBP and SBP), and original body mass index (BMI)), and lab tests (i.e., Logical Observation Identifiers, Names, and Codes (LOINC)). Missing values for DBP and SBP were imputed using their respective empirical means computed from the observed data. The two steps required for processing medication data are as follows: (1) NDC codes were converted to RXCUIs, and (2) RXCUIs were further mapped to ingredient-level RXCUIs to reduce redundancy. All features were selected according to their clinical relevance to HF and availability in routine care settings [20, 21]. Additionally, we incorporated relative time information, defined as the difference in days between clinical encounters, to capture the dynamic trajectories of patients [17]. By integrating a variety of relevant features, this study aims to improve the model’s ability to identify patterns indicative of HF, thus supporting timely and accurate risk stratification.

### 2.3. Definitions of cases and controls

To construct a matched control cohort, we started with an initial cohort of 4,503,509 patients from the OneFlorida dataset. We defined the date of HF onset as the date of the first encounter of three consecutive HF diagnosis encounters that occurred within 12 months. In particular, HF cases were identified according to the following inclusion criteria: (i) patients must have had at least three clinical encounters coded with qualified HF diagnoses based on ICD-10 codes (as shown in Table 1) within a 12-month window [20]; (ii) patients must be 60 years of age or older at the time of the first HF-related encounter to focus on a high-risk population [19]; and (iii) patients must have at least one medication record indicating the use of guideline-directed medical therapy for HF, which includes Angiotensin-Converting Enzyme Inhibitor, Beta-Blocker, Angiotensin II Receptor Blockers, and Loop Diuretic [22].

**Table 1:**
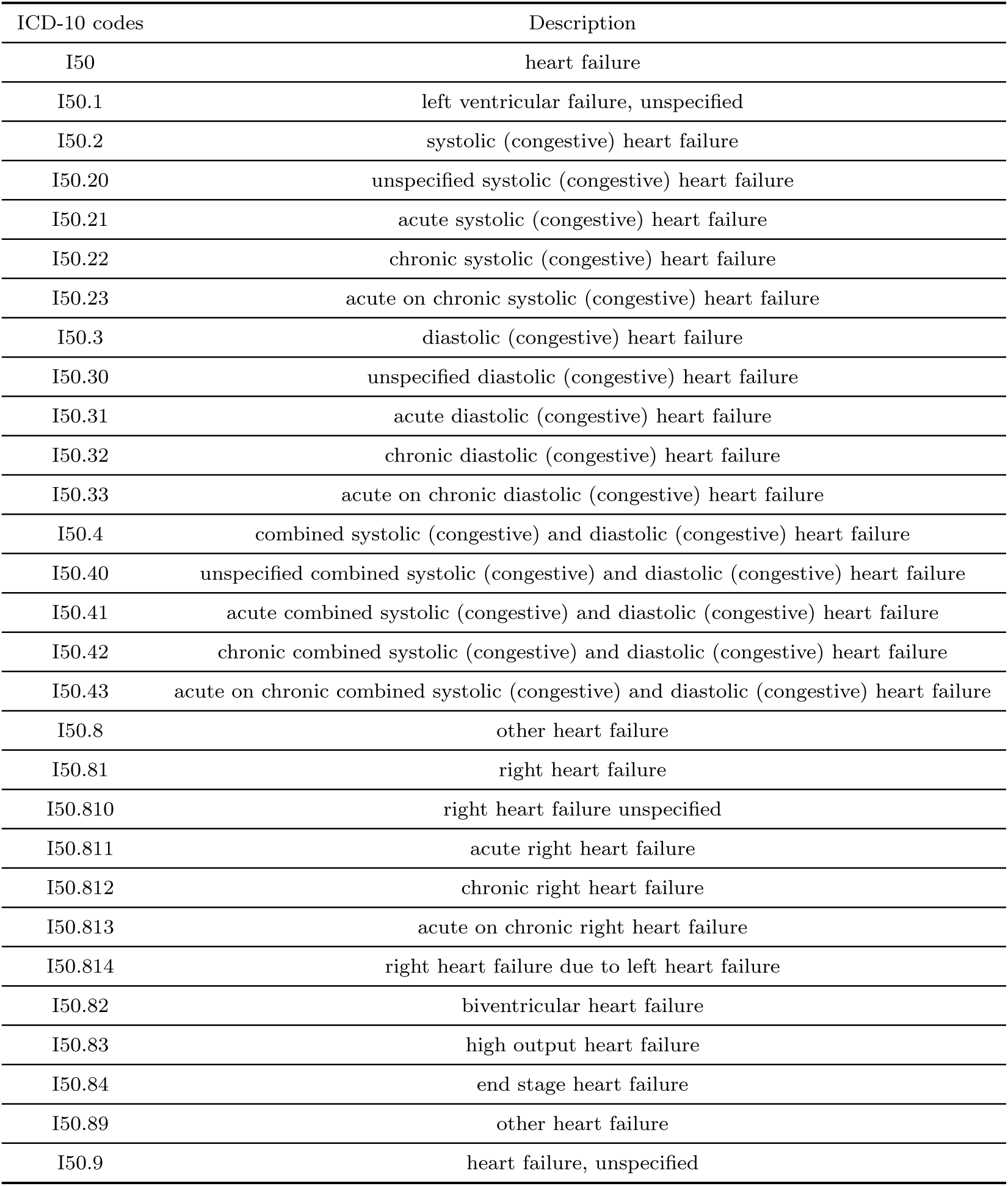
ICD-10 codes for HF diagnosis.

| ICD-10 codes | Description |
| --- | --- |
| I50 | heart failure |
| I50.1 | left ventricular failure, unspecified |
| I50.2 | systolic (congestive) heart failure |
| I50.20 | unspecified systolic (congestive) heart failure |
| I50.21 | acute systolic (congestive) heart failure |
| I50.22 | chronic systolic (congestive) heart failure |
| I50.23 | acute on chronic systolic (congestive) heart failure |
| I50.3 | diastolic (congestive) heart failure |
| I50.30 | unspecified diastolic (congestive) heart failure |
| I50.31 | acute diastolic (congestive) heart failure |
| I50.32 | chronic diastolic (congestive) heart failure |
| I50.33 | acute on chronic diastolic (congestive) heart failure |
| I50.4 | combined systolic (congestive) and diastolic (congestive) heart failure |
| I50.40 | unspecified combined systolic (congestive) and diastolic (congestive) heart failure |
| I50.41 | acute combined systolic (congestive) and diastolic (congestive) heart failure |
| I50.42 | chronic combined systolic (congestive) and diastolic (congestive) heart failure |
| I50.43 | acute on chronic combined systolic (congestive) and diastolic (congestive) heart failure |
| I50.8 | other heart failure |
| I50.81 | right heart failure |
| I50.810 | right heart failure unspecified |
| I50.811 | acute right heart failure |
| I50.812 | chronic right heart failure |
| I50.813 | acute on chronic right heart failure |
| I50.814 | right heart failure due to left heart failure |
| I50.82 | biventricular heart failure |
| I50.83 | high output heart failure |
| I50.84 | end stage heart failure |
| I50.89 | other heart failure |
| I50.9 | heart failure, unspecified |

The objective of matching cases (patients with HF) to controls (patients without HF) is to ensure that both groups are comparable and have similar observation durations. By doing this, any differences in results can be attributed to the medical condition (HF) rather than to differences in the availability of data. For each case, controls were identified according to the following inclusion criteria: (i) matching encounter type, gender, and race with the corresponding case to account for potential confounding demographics [19]; (ii) age within a five-year range of the case to account for age-related risk factors [19]; (iii) the first encounter of the control must have occurred within 365 days of the case’s first encounter, which ensures that both patients entered the healthcare system within a similar timeframe, minimizing bias due to discrepancies in medical record availability; and (iv) at least one clinical encounter must have occurred either within 30 days prior to or at any point after the case’s HF onset date, ensuring that the control was actively engaged in the healthcare system during the period of interest.

A note of caution is due here: for each control, the encounter date closest to the HF onset date of the corresponding case was used as the reference encounter date to align the timeline across both. Cases that could not be matched to at least one eligible control based on the criteria above were excluded from the dataset. Together, a total of 2561 cases of HF and 5493 controls were identified, as shown in Table 2. This rigorous case-control matching strategy enhances the validity of downstream analysis by controlling for demographic and temporal confounders while minimizing selection bias.

**Table 2:** Baseline characteristics of the case and control cohorts.

| Variables | Sub Types | Case (n=2,561) | Control (n=5,493) |
| --- | --- | --- | --- |
| Age | 60-64 | 6.65% | 7.60% |
|  | 65-69 | 23.88% | 25.97% |
|  | 70-74 | 24.96% | 25.94% |
|  | 75-79 | 18.98% | 17.09% |
|  | 80-84 | 14.53% | 12.88% |
| | $\geq 85$ | 11.00% | 10.52% |
| Gender | Male | 48.14% | 49.84% |
|  | Female | 51.86% | 50.16% |
| Race | White | 63.32% | 64.12% |
|  | Black | 30.79% | 30.16% |
|  | Asian | 3.16% | 3.25% |
|  | Other | 2.73% | 2.47% |
| Ethnicity | Hispanic | 2.04% | 1.95% |
|  | Non-Hispanic | 93.79% | 93.92% |
|  | Other | 4.17% | 4.13% |
| Vital signs | Diastolic blood pressure | 75.92 | 76.46 |
|  | Systolic blood pressure | 139.97 | 138.32 |
|  | Original body mass index | 30.56 | 28.98 |

### 2.4. Observation period and prediction window

The utility of a predictive model depends not only on its accuracy but also on the availability of input data and the time frame in which it can provide reliable predictions. In this study, we define two critical temporal parameters: the observation window and the prediction window. The former refers to the period during which patient data is collected, and the latter refers to how far into the future the model aims to predict the outcomes. For clinical purposes, we have adopted a 12-month observation window to capture the patient’s medical history and a 6-month prediction window to facilitate timely identification of HF-related outcomes [20, 21]. This prediction horizon is particularly useful for studying early intervention and proactive disease management [12]. It should be noted that extending the prediction window could introduce additional uncertainty due to possible changes in patient health status, treatment plans, or healthcare utilization over time. These temporal dynamics should be carefully taken into account when interpreting model performance.

### 2.5. Model development

In order to generate dense vector representations from structured medical codes, we explored three NLP-based approaches: BoW, Skip-gram, and ClinicalBERT, as shown in Figure. We applied these three embedding methods to four types of medical codes: ICD-10, CPT, ingredient-level RXCUIs, and LOINC. The BoW is a frequency-based encoding method that treats each unique medical code as a token. Patient representations are constructed by counting the occurrences of these codes, resulting in sparse vectors that are subsequently projected into dense embeddings. The Skip-gram [23] is a predictive embedding approach using the Word2Vec framework. It learns distributed representations by maximizing the likelihood of context codes given a target code. This method captures local co-occurrence patterns among medical codes. We trained separate Skip-gram models for each type of medical code to obtain their dense embeddings. The ClinicalBERT [16] is a transformer-based language model pre-trained on large-scale clinical corpora. We fine-tuned ClinicalBERT using labeled data in a supervised manner. For each patient, contextualized embeddings were extracted from the final hidden layer and then used as input features. Moreover, patient demographics and vital signs were directly concatenated with the embeddings, creating a final feature matrix for downstream classification.

The two baseline models were used in this study. GRU [24] is a type of recurrent neural network that models multiple encounters as temporal sequences. The input data for the GRU is derived from embeddings generated by the skip-gram model and processed through a single-layer GRU to capture longitudinal patterns, as used in the study [20]. The final hidden representation of the GRU is then passed through a dropout layer followed by a fully connected layer for downstream classification. RETAIN [25] is a recurrent neural network model designed to improve the modeling of EHR data by explicitly learning attention weights in both encounters and variables. It employs two gated recurrent units that compute attention weights for encounters and variables. These attention weights are combined to create a context vector fed into a classifier for downstream classification. Similar to GRU, RETAIN also uses embeddings generated by the skip-gram model as input. The modeling of patient demographics and vital signs follows the same step as previously described.

To assess the performance of the model, we conducted a 10-fold cross-validation on the OneFlorida dataset, as illustrated in Figure 1. The dataset was divided into ten equal-sized, nonoverlapping subsets. In each fold, one subset served as the validation set, while the remaining nine were combined to establish the training set. We trained five prediction models, including Logistic Regression (LR), Lasso, Random Forest (RF), XGBoost, and Multilayer Perceptron (MLP), on the training data and evaluated their performance on the held-out validation set. This process was repeated ten times, with each subset being used once as the validation set, providing a robust performance assessment. The same assessment was applied to both baseline models to ensure a fair comparison.

**Figure 1:**
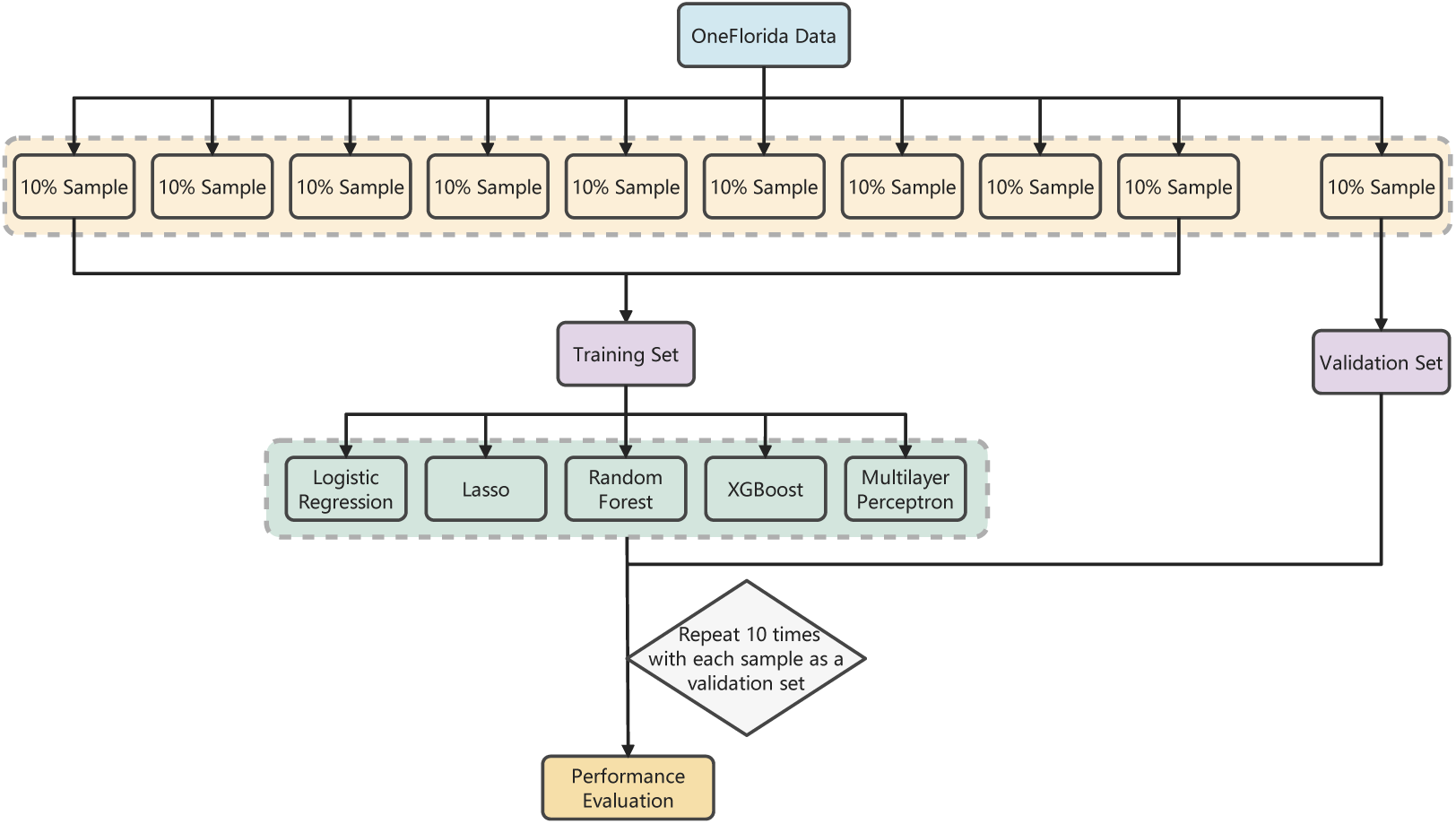
Model Evaluation Workflow

### 2.6. Performance metrics

To comprehensively assess model performance, various evaluation metrics are employed, including the Area Under the Receiver Operating Characteristic Curve (AUROC), Precision, Recall, Accuracy, F1 Score, the Area Under the Precision-Recall Curve (AUPRC), Matthews Correlation Coefficient (MCC), and Brier Score. Specifically, AUROC evaluates the model’s ability to distinguish between classes across thresholds and is effective for balanced settings. Precision measures the proportion of correct positive predictions, while Recall assesses the model’s ability to identify actual positive instances. Accuracy reflects the overall correctness of the model but can be misleading with imbalanced settings. F1 Score makes a tradeoff between Precision and Recall, making it useful when the costs of false positives and false negatives are similar. AUPRC emphasizes the positive class and is often more informative than AUROC in imbalanced settings. MCC captures the overall correlation between predicted and actual labels, making it particularly useful for binary classification tasks with skewed class distributions. Finally, the Brier Score measures the accuracy of probabilistic predictions, taking into account both calibration and discrimination.

## 3. Results

### 3.1. Results of early HF onset detection

Table 3 provides a comprehensive performance comparison of the baseline models for the early detection of HF onset in a variety of evaluation metrics. These results suggest that tree-based models, including XGBoost and RF, consistently outperform other models in most evaluation metrics. Specifically, XGBoost achieved the highest performance across key metrics, with an AUROC of 0.7672 ± 0.0156, an F1 score of 0.5547 ± 0.0308, an AUPRC of 0.6380 ± 0.0183, and a MCC of 0.3983 ± 0.0386, indicating its strong discriminative power and overall robustness. RF achieved competitive performance, with an AUROC of 0.7556 ± 0.0223, a precision of 0.6905 ± 0.0313, and an F1 score of 0.5173 ± 0.0254. LR and Lasso provided moderate and balanced performance in most metrics. Lasso improved over LR in precision and AUROC, likely due to its focus on feature sparsity. In contrast, neural network models, including MLP, GRU, and RETAIN, demonstrated lower performance across most metrics. While MLP achieved a competitive recall of 0.5057 ± 0.0428, its suboptimal precision and F1 score suggest oversensitivity, leading to increased false positives. GRU and RETAIN presented lower performance, with recall values of 0.1109 and 0.1777, respectively, and F1 scores of 0.1829 and 0.2628, respectively. Overall, these results highlight the superiority of tree-based models over neural network models for early detection of HF onset in the given experimental setting.

**Table 3:** Performance comparison of baseline models on early detection of HF onset.

| Models | Metrics |  |  |  |
| --- | --- | --- | --- | --- |
|  | AUROC | Precision | Recall | Accuracy |
| LR | 0.7145 ± 0.0189 | 0.5643 ± 0.0295 | 0.4959 ± 0.0366 | 0.7175 ± 0.0149 |
| Lasso | 0.7298 ± 0.0199 | 0.5968 ± 0.0185 | 0.4983 ± 0.0401 | 0.7312 ± 0.0096 |
| RF | 0.7556 ± 0.0223 | 0.6905 ± 0.0313 | 0.4139 ± 0.0248 | 0.7545 ± 0.0121 |
| XGBoost | 0.7672 ± 0.0156 | 0.6542 ± 0.0331 | 0.4822 ± 0.0342 | 0.7542 ± 0.0150 |
| MLP | 0.6972 ± 0.0209 | 0.5093 ± 0.0217 | 0.5057 ± 0.0482 | 0.6870 ± 0.0138 |
| GRU | 0.6496 ± 0.0168 | 0.5604 ± 0.0560 | 0.1109 ± 0.0299 | 0.6888 ± 0.0048 |
| RETAIN | 0.6616 ± 0.0206 | 0.5498 ± 0.0341 | 0.1777 ± 0.0639 | 0.6923 ± 0.0077 |
|  | F1 Score | AUPRC | MCC | Brier score |
| LR | 0.5272 ± 0.0267 | 0.5619 ± 0.0244 | 0.3290 ± 0.0350 | 0.2162 ± 0.0093 |
| Lasso | 0.5423 ± 0.0262 | 0.5873 ± 0.0290 | 0.3598 ± 0.0279 | 0.1999 ± 0.0084 |
| RF | 0.5173 ± 0.0254 | 0.6259 ± 0.0318 | 0.3880 ± 0.0324 | 0.1740 ± 0.0066 |
| XGBoost | 0.5547 ± 0.0308 | 0.6382 ± 0.0188 | 0.3993 ± 0.0388 | 0.1875 ± 0.0092 |
| MLP | 0.5060 ± 0.0223 | 0.5515 ± 0.0320 | 0.2784 ± 0.0250 | 0.2759 ± 0.0131 |
| GRU | 0.1829 ± 0.0407 | 0.4500 ± 0.0224 | 0.1323 ± 0.0253 | 0.2047 ± 0.0029 |
| RETAIN | 0.2628 ± 0.0698 | 0.4677 ± 0.0276 | 0.1668 ± 0.0408 | 0.2027 ± 0.0038 |

### 3.2. Analysis on feature importance and absolute coefficients

Table 4 presents a detailed analysis of feature importance and absolute coefficients in four state-of-the-art models. Diagnoses/ICD-10 codes consistently demonstrated the highest predictive contributions, especially in linear models. Notably, LR and Lasso demonstrated significantly higher absolute coefficients for ICD-10 embeddings, with values of 15.56 for LR and 11.97 for Lasso when using embeddings generated from BoW. Among demographic and physiological variables, age and race are important features within linear models, indicating their relevance in risk stratification. Consistent with our expectations, Lasso encouraged model sparsity, resulting in fewer high-magnitude coefficients compared to LR, which could improve interpretability in clinical settings. The embeddings generated from ClinicalBERT outperformed those from the BoW and Skip-gram models when applied to medications/RXCUIs and lab tests/LOINC codes, especially in linear models. Tree-based models include RF and XGBoost, showing more evenly distributed importance scores but still prioritizing ICD-10 codes. For example, XGBoost assigned the highest importance to ICD-10 embeddings, with a total sum of 0.0754, mainly due to embeddings generated from ClinicalBERT, which contribute a feature importance of 0.0752.

**Table 4:**
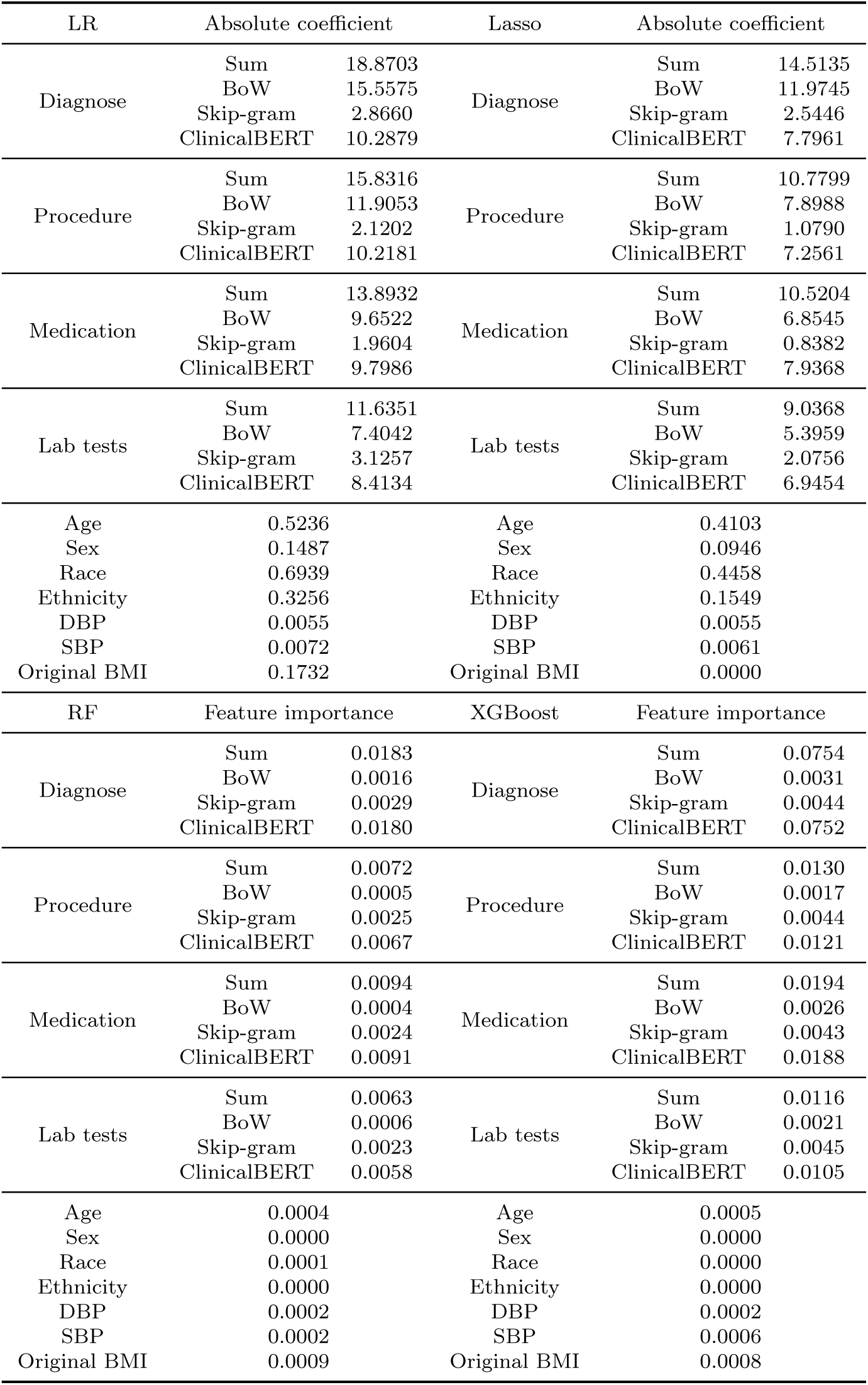
Analysis of feature importance and absolute coefficients across embedding methods and feature types.

| LR | Absolute coefficient |  | Lasso | Absolute coefficient |  |
| --- | --- | --- | --- | --- | --- |
| Diagnose | Sum | 18.8703 | Diagnose | Sum | 14.5135 |
|  | BoW | 15.5575 |  | BoW | 11.9745 |
|  | Skip-gram | 2.8660 |  | Skip-gram | 2.5446 |
|  | ClinicalBERT | 10.2879 |  | ClinicalBERT | 7.7961 |
| Procedure | Sum | 15.8316 | Procedure | Sum | 10.7799 |
|  | BoW | 11.9053 |  | BoW | 7.8988 |
|  | Skip-gram | 2.1202 |  | Skip-gram | 1.0790 |
|  | ClinicalBERT | 10.2181 |  | ClinicalBERT | 7.2561 |
| Medication | Sum | 13.8932 | Medication | Sum | 10.5204 |
|  | BoW | 9.6522 |  | BoW | 6.8545 |
|  | Skip-gram | 1.9604 |  | Skip-gram | 0.8382 |
|  | ClinicalBERT | 9.7986 |  | ClinicalBERT | 7.9368 |
| Lab tests | Sum | 11.6351 | Lab tests | Sum | 9.0368 |
|  | BoW | 7.4042 |  | BoW | 5.3959 |
|  | Skip-gram | 3.1257 |  | Skip-gram | 2.0756 |
|  | ClinicalBERT | 8.4134 |  | ClinicalBERT | 6.9454 |
| Age | 0.5236 |  | Age | 0.4103 |  |
| Sex | 0.1487 |  | Sex | 0.0946 |  |
| Race | 0.6939 |  | Race | 0.4458 |  |
| Ethnicity | 0.3256 |  | Ethnicity | 0.1549 |  |
| DBP | 0.0055 |  | DBP | 0.0055 |  |
| SBP | 0.0072 |  | SBP | 0.0061 |  |
| Original BMI | 0.1732 |  | Original BMI | 0.0000 |  |
| RF | Feature importance |  | XGBoost | Feature importance |  |
| Diagnose | Sum | 0.0183 | Diagnose | Sum | 0.0754 |
|  | BoW | 0.0016 |  | BoW | 0.0031 |
|  | Skip-gram | 0.0029 |  | Skip-gram | 0.0044 |
|  | ClinicalBERT | 0.0180 |  | ClinicalBERT | 0.0752 |
| Procedure | Sum | 0.0072 | Procedure | Sum | 0.0130 |
|  | BoW | 0.0005 |  | BoW | 0.0017 |
|  | Skip-gram | 0.0025 |  | Skip-gram | 0.0044 |
|  | ClinicalBERT | 0.0067 |  | ClinicalBERT | 0.0121 |
| Medication | Sum | 0.0094 | Medication | Sum | 0.0194 |
|  | BoW | 0.0004 |  | BoW | 0.0026 |
|  | Skip-gram | 0.0024 |  | Skip-gram | 0.0043 |
|  | ClinicalBERT | 0.0091 |  | ClinicalBERT | 0.0188 |
| Lab tests | Sum | 0.0063 | Lab tests | Sum | 0.0116 |
|  | BoW | 0.0006 |  | BoW | 0.0021 |
|  | Skip-gram | 0.0023 |  | Skip-gram | 0.0045 |
|  | ClinicalBERT | 0.0058 |  | ClinicalBERT | 0.0105 |
| Age | 0.0004 |  | Age | 0.0005 |  |
| Sex | 0.0000 |  | Sex | 0.0000 |  |
| Race | 0.0001 |  | Race | 0.0000 |  |
| Ethnicity | 0.0000 |  | Ethnicity | 0.0000 |  |
| DBP | 0.0002 |  | DBP | 0.0002 |  |
| SBP | 0.0002 |  | SBP | 0.0006 |  |
| Original BMI | 0.0009 |  | Original BMI | 0.0008 |  |

### 3.3. Results of early HF onset detection across gender, race, and ethnicity subgroups

Tables 5, 6, and 7 compare the performance of early detection of HF onset across gender, race, and ethnicity subgroups using baseline models. As Table 5 shows, Lasso consistently outperformed LR in most subgroups and evaluation metrics. This superior performance was demonstrated by a higher AUROC, improved calibration indicated by lower Brier scores, and enhanced predictive power showed by higher F1 scores and MCC. The performance between male and female subgroups is comparable across both models, although male subgroups achieved slightly higher AUROC and accuracy when using Lasso.

**Table 5:** Comparative analysis of early HF onset detection performance across gender, race, and ethnicity subgroups using linear models.

| LR | AUROC | Precision | Recall | Accuracy |
| --- | --- | --- | --- | --- |
| Male | 0.7220 ± 0.0246 | 0.5591 ± 0.0573 | 0.5095 ± 0.0405 | 0.7168 ± 0.0181 |
| Female | 0.7100 ± 0.0209 | 0.5714 ± 0.0312 | 0.5031 ± 0.0442 | 0.7203 ± 0.0170 |
| White | 0.7133 ± 0.0267 | 0.5664 ± 0.0379 | 0.5146 ± 0.0460 | 0.7153 ± 0.0165 |
| Black | 0.7256 ± 0.0395 | 0.5639 ± 0.0461 | 0.4999 ± 0.0570 | 0.7281 ± 0.0282 |
| Asian | 0.7264 ± 0.1385 | 0.5478 ± 0.2400 | 0.4961 ± 0.2406 | 0.7304 ± 0.1058 |
| Hispanic | 0.7675 ± 0.1102 | 0.6068 ± 0.1428 | 0.4959 ± 0.1813 | 0.7392 ± 0.1003 |
| Non-Hispanic | 0.7163 ± 0.0196 | 0.5663 ± 0.0278 | 0.5097 ± 0.0285 | 0.7197 ± 0.0134 |
| LR | F1 Score | AUPRC | MCC | Brier score |
| Male | 0.5315 ± 0.0370 | 0.5695 ± 0.0599 | 0.3316 ± 0.0496 | 0.2174 ± 0.0146 |
| Female | 0.5337 ± 0.0297 | 0.5668 ± 0.0177 | 0.3378 ± 0.0371 | 0.2192 ± 0.0121 |
| White | 0.5378 ± 0.0323 | 0.5726 ± 0.0426 | 0.3351 ± 0.0416 | 0.2206 ± 0.0140 |
| Black | 0.5291 ± 0.0476 | 0.5653 ± 0.0569 | 0.3408 ± 0.0639 | 0.2122 ± 0.0231 |
| Asian | 0.5066 ± 0.2288 | 0.6467 ± 0.1124 | 0.3317 ± 0.2298 | 0.2058 ± 0.0782 |
| Hispanic | 0.5357 ± 0.1586 | 0.7063 ± 0.1211 | 0.3650 ± 0.2081 | 0.1917 ± 0.0751 |
| Non-Hispanic | 0.5362 ± 0.0248 | 0.5649 ± 0.0323 | 0.3374 ± 0.0328 | 0.2177 ± 0.0100 |
| Lasso | AUROC | Precision | Recall | Accuracy |
| Male | 0.7317 ± 0.0271 | 0.5858 ± 0.0582 | 0.4967 ± 0.0216 | 0.7316 ± 0.0162 |
| Female | 0.7257 ± 0.0204 | 0.5923 ± 0.0297 | 0.4956 ± 0.0315 | 0.7309 ± 0.0176 |
| White | 0.7232 ± 0.0278 | 0.5927 ± 0.0369 | 0.4983 ± 0.0355 | 0.7269 ± 0.0172 |
| Black | 0.7427 ± 0.0430 | 0.5841 ± 0.0411 | 0.5037 ± 0.0651 | 0.7381 ± 0.0272 |
| Asian | 0.7484 ± 0.1515 | 0.5650 ± 0.2482 | 0.4767 ± 0.2570 | 0.7424 ± 0.1382 |
| Hispanic | 0.7914 ± 0.0888 | 0.6350 ± 0.1829 | 0.4809 ± 0.2027 | 0.7526 ± 0.0924 |
| Non-Hispanic | 0.7288 ± 0.0198 | 0.5900 ± 0.0251 | 0.4994 ± 0.0162 | 0.7301 ± 0.0115 |
| Lasso | F1 Score | AUPRC | MCC | Brier score |
| Male | 0.5366 ± 0.0312 | 0.5840 ± 0.0550 | 0.3503 ± 0.0456 | 0.2009 ± 0.0124 |
| Female | 0.5389 ± 0.0241 | 0.5851 ± 0.0195 | 0.3532 ± 0.0337 | 0.2025 ± 0.0116 |
| White | 0.5406 ± 0.0288 | 0.5847 ± 0.0430 | 0.3519 ± 0.0410 | 0.2050 ± 0.0139 |
| Black | 0.5397 ± 0.0502 | 0.5861 ± 0.0549 | 0.3612 ± 0.0639 | 0.1943 ± 0.0223 |
| Asian | 0.5066 ± 0.2477 | 0.6810 ± 0.1337 | 0.3406 ± 0.2850 | 0.1815 ± 0.0809 |
| Hispanic | 0.5362 ± 0.1919 | 0.7200 ± 0.0990 | 0.3846 ± 0.2240 | 0.1800 ± 0.0637 |
| Non-Hispanic | 0.5407 ± 0.0172 | 0.5813 ± 0.0305 | 0.3541 ± 0.0260 | 0.2012 ± 0.0086 |

**Table 6:**
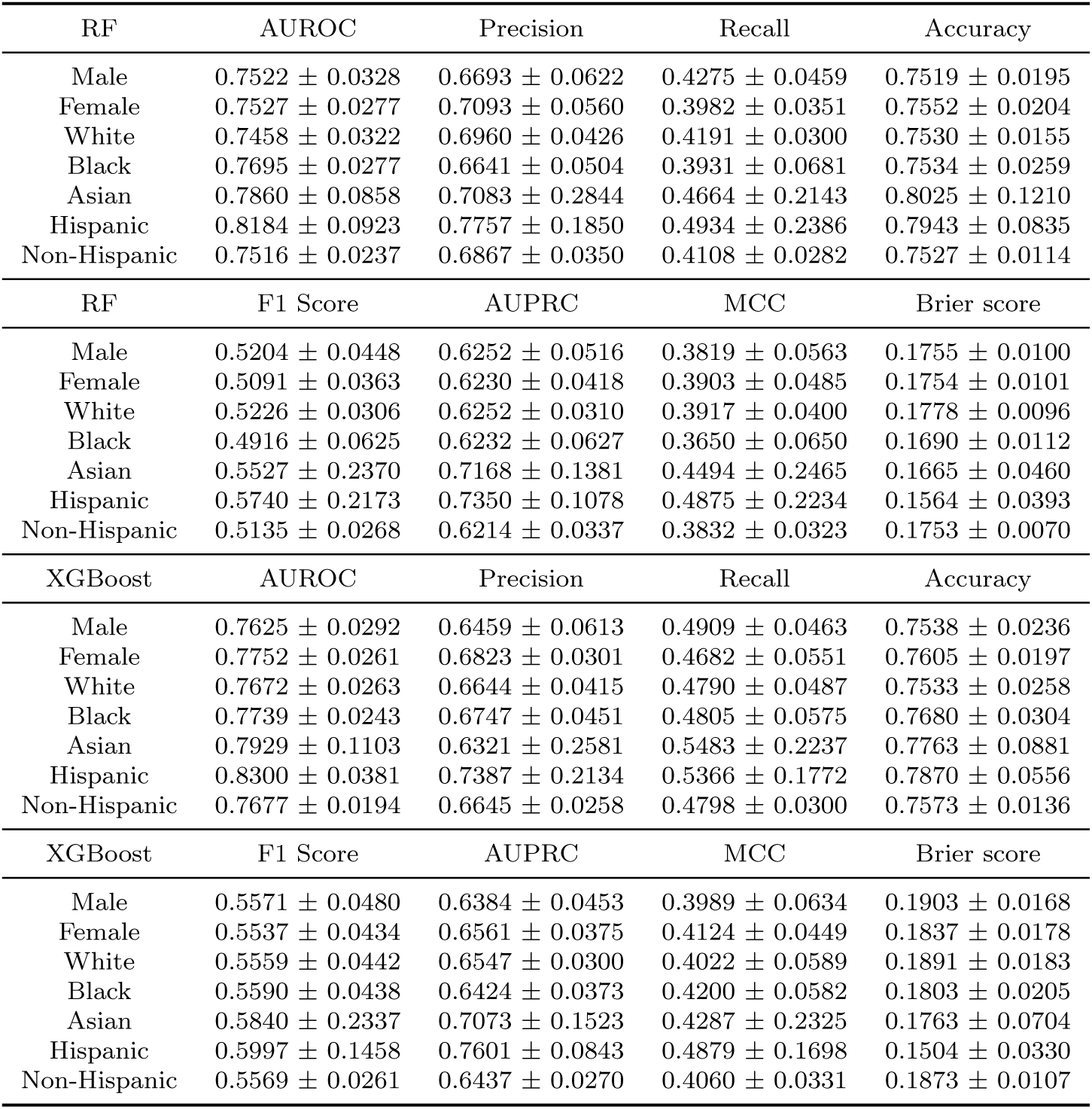
Comparative analysis of early HF onset detection performance across gender, race, and ethnicity subgroups using tree-based models.

Significant differences were observed between the race and ethnicity subgroups. In particular, the Hispanic subgroup demonstrated superior performance with Lasso, achieving an AUROC of 0.7914 ± 0.0888, an AUPRC of 0.7200 ± 0.0990, and an MCC of 0.3846 ± 0.2240. The standard deviations associated with these metrics indicate variability in model performance, with Asian and Hispanic subgroups showing greater variance. This variability may be attributed to data imbalance or underlying subgroup-specific heterogeneity, which can affect the generalizability of the model.

As can be seen from Table 6, XGBoost outperformed RF in most subgroups and key evaluation metrics with AUROC, AUPRC, and F1 scores. When comparing performance by gender, male subgroups outperformed female subgroups in terms of recall and F1 score for both models. For example, the male subgroup using RF achieved a recall of 0.4275 ± 0.0459 compared to 0.3982 ± 0.0351 and an F1 score of 0.5204 ± 0.0448 compared to 0.5091 ± 0.0363 for female subgroups. Among the ethnicity subgroups, the Hispanic group using XGBoost demonstrated strong performance, achieving the highest AUROC of 0.8300 ± 0.0381, AUPRC of 0.7601 ± 0.0843, and MCC of 0.4879 ± 0.1698 across all subgroups. These results indicated accurate and well-calibrated predictions in this subgroup. Notably, the Black subgroup showed lower performance, especially with RF, which yields the lowest F1 score of 0.4916 ± 0.0265) and an MCC of 0.3650 ± 0.0650. Finally, MLP consistently outperformed GRU and RETAIN across all evaluation metrics, as shown in Table 7. This improved performance was evident across gender, race, and ethnicity subgroups.

**Table 7:** Comparative analysis of early HF onset detection performance across gender, race, and ethnicity subgroups using neural network models.

| MLP | AUROC | Precision | Recall | Accuracy |
| --- | --- | --- | --- | --- |
| Male | 0.7010 ± 0.0283 | 0.5307 ± 0.0572 | 0.4996 ± 0.0448 | 0.7007 ± 0.0208 |
| Female | 0.7058 ± 0.0239 | 0.5413 ± 0.0357 | 0.5029 ± 0.0413 | 0.7045 ± 0.0213 |
| White | 0.7012 ± 0.0287 | 0.5430 ± 0.0402 | 0.5026 ± 0.0378 | 0.7011 ± 0.0196 |
| Black | 0.7059 ± 0.0355 | 0.5274 ± 0.0396 | 0.5025 ± 0.0626 | 0.7101 ± 0.0266 |
| Asian | 0.6949 ± 0.1706 | 0.5054 ± 0.3084 | 0.4353 ± 0.2576 | 0.7003 ± 0.1042 |
| Hispanic | 0.7506 ± 0.1139 | 0.6375 ± 0.1716 | 0.5016 ± 0.1806 | 0.7440 ± 0.1109 |
| Non-Hispanic | 0.7021 ± 0.0194 | 0.5358 ± 0.0284 | 0.5018 ± 0.0300 | 0.7025 ± 0.0171 |
| MLP | F1 Score | AUPRC | MCC | Brier score |
| Male | 0.5128 ± 0.0387 | 0.5488 ± 0.0484 | 0.2994 ± 0.0535 | 0.2644 ± 0.0180 |
| Female | 0.5202 ± 0.0302 | 0.5656 ± 0.0297 | 0.3085 ± 0.0416 | 0.2588 ± 0.0225 |
| White | 0.5203 ± 0.0249 | 0.5569 ± 0.0361 | 0.3058 ± 0.0383 | 0.2637 ± 0.0165 |
| Black | 0.5137 ± 0.0496 | 0.5539 ± 0.0470 | 0.3076 ± 0.0592 | 0.2547 ± 0.0309 |
| Asian | 0.4552 ± 0.2556 | 0.6307 ± 0.1914 | 0.2654 ± 0.2654 | 0.2611 ± 0.1042 |
| Hispanic | 0.5489 ± 0.1760 | 0.6886 ± 0.1210 | 0.3815 ± 0.2302 | 0.2204 ± 0.0921 |
| Non-Hispanic | 0.5176 ± 0.0231 | 0.5524 ± 0.0295 | 0.3039 ± 0.0351 | 0.2619 ± 0.0175 |
| GRU | AUROC | Precision | Recall | Accuracy |
| Male | 0.6557 ± 0.0097 | 0.5437 ± 0.0732 | 0.1323 ± 0.0438 | 0.6900 ± 0.0168 |
| Female | 0.6473 ± 0.0304 | 0.5914 ± 0.0901 | 0.0897 ± 0.0238 | 0.6875 ± 0.0166 |
| White | 0.6524 ± 0.0182 | 0.5682 ± 0.0580 | 0.1189 ± 0.0273 | 0.6853 ± 0.0135 |
| Black | 0.6363 ± 0.0355 | 0.5655 ± 0.1419 | 0.1018 ± 0.0414 | 0.6982 ± 0.0200 |
| Asian | 0.7476 ± 0.0754 | 0.2963 ± 0.4287 | 0.1049 ± 0.1746 | 0.7070 ± 0.1359 |
| Hispanic | 0.7000 ± 0.0716 | 0.3500 ± 0.3905 | 0.0657 ± 0.0784 | 0.6803 ± 0.0659 |
| Non-Hispanic | 0.6498 ± 0.0207 | 0.5677 ± 0.0643 | 0.1139 ± 0.0317 | 0.6895 ± 0.0070 |
| GRU | F1 Score | AUPRC | MCC | Brier score |
| Male | 0.2094 ± 0.0570 | 0.4543 ± 0.0400 | 0.1386 ± 0.0425 | 0.2031 ± 0.0056 |
| Female | 0.1535 ± 0.0357 | 0.4553 ± 0.0480 | 0.1273 ± 0.0287 | 0.2063 ± 0.0071 |
| White | 0.1947 ± 0.0366 | 0.4648 ± 0.0229 | 0.1386 ± 0.0276 | 0.2055 ± 0.0060 |
| Black | 0.1683 ± 0.0566 | 0.4321 ± 0.0608 | 0.1300 ± 0.0583 | 0.2035 ± 0.0055 |
| Asian | 0.1457 ± 0.2336 | 0.6554 ± 0.1445 | 0.1176 ± 0.2259 | 0.1914 ± 0.0514 |
| Hispanic | 0.1036 ± 0.1133 | 0.5307 ± 0.0810 | 0.0595 ± 0.1437 | 0.1977 ± 0.0323 |
| Non-Hispanic | 0.1872 ± 0.0424 | 0.4522 ± 0.0223 | 0.1369 ± 0.0276 | 0.2047 ± 0.0048 |
| RETAIN | AUROC | Precision | Recall | Accuracy |
| Male | 0.6688 ± 0.0159 | 0.5482 ± 0.0549 | 0.2125 ± 0.0790 | 0.6967 ± 0.0109 |
| Female | 0.6578 ± 0.0348 | 0.5525 ± 0.0749 | 0.1427 ± 0.0524 | 0.6879 ± 0.0179 |
| White | 0.6652 ± 0.0206 | 0.5744 ± 0.0430 | 0.1954 ± 0.0621 | 0.6928 ± 0.0144 |
| Black | 0.6479 ± 0.0370 | 0.5008 ± 0.0931 | 0.1539 ± 0.0745 | 0.6945 ± 0.0266 |
| Asian | 0.7504 ± 0.0775 | 0.4630 ± 0.3831 | 0.1849 ± 0.1692 | 0.7087 ± 0.1333 |
| Hispanic | 0.7110 ± 0.0672 | 0.4000 ± 0.4359 | 0.0723 ± 0.0777 | 0.6833 ± 0.0613 |
| Non-Hispanic | 0.6627 ± 0.0232 | 0.5546 ± 0.0309 | 0.1827 ± 0.0694 | 0.6934 ± 0.0106 |
| RETAIN | F1 Score | AUPRC | MCC | Brier score |
| Male | 0.2990 ± 0.0834 | 0.4762 ± 0.0467 | 0.1858 ± 0.0524 | 0.2008 ± 0.0055 |
| Female | 0.2216 ± 0.0617 | 0.4694 ± 0.0431 | 0.1466 ± 0.0440 | 0.2045 ± 0.0089 |
| White | 0.2861 ± 0.0648 | 0.4829 ± 0.0290 | 0.1867 ± 0.0411 | 0.2033 ± 0.0059 |
| Black | 0.2286 ± 0.0901 | 0.4470 ± 0.0645 | 0.1355 ± 0.0770 | 0.2019 ± 0.0060 |
| Asian | 0.2504 ± 0.2133 | 0.6340 ± 0.0652 | 0.1653 ± 0.2209 | 0.1891 ± 0.0558 |
| Hispanic | 0.1189 ± 0.1257 | 0.5522 ± 0.1228 | 0.0633 ± 0.1774 | 0.1927 ± 0.0285 |
| Non-Hispanic | 0.2682 ± 0.0745 | 0.4704 ± 0.0264 | 0.1722 ± 0.0444 | 0.2027 ± 0.0053 |

### 3.4. Model calibration

We evaluated the calibration performance of LR, Lasso, and MLP, as shown in Table 8. This evaluation was conducted under three calibration scenarios: no recalibration, recalibration in the large, and logistic recalibration. Model calibration was measured following the calibration hierarchy, using weak calibration metrics (intercept and slope) and moderate calibration metrics (the Brier score, Emax, and Eavg as detailed by [26]). The ideal values for the intercept and slope are 0 and 1, respectively. The Brier score measures the mean squared error between predicted probabilities and actual outcomes, where lower values indicate better performance. Emax and Eavg represent the maximum and average deviation between predicted probabilities and observed outcomes, with lower values indicating better calibration. All estimations included 95% confidence intervals, which were derived using bootstrapping techniques.

**Table 8:** Overall calibration statistics for LR, Lasso, and MLP.

| Methods | Calibration Metric |  |
| --- | --- | --- |
| No Recalibration (95\% CI) | Calibration Intercept | Calibration Slope |
| LR | -0.5298 [-0.5722, -0.4874] | 0.2201 [0.1904, 0.2497] |
| Lasso | -0.4741 [-0.5178, -0.4304] | 0.3106 [0.2716, 0.3497] |
| MLP | -0.5889 [-0.6310, -0.5467] | 0.0754 [0.0634, 0.0874] |
| Recalibration In the Large (95\% CI) | Calibration Intercept | Calibration Slope |
| LR | -0.5245 [-0.5504, -0.4986] | 0.2201 [0.1904, 0.2497] |
| Lasso | -0.4670 [-0.4981, -0.4358] | 0.3106 [0.2716, 0.3497] |
| MLP | -0.5784 [-0.5986, -0.5582] | 0.0754 [0.0634, 0.0874] |
| Logistic Recalibration (95\% CI) | Calibration Intercept | Calibration Slope |
| LR | 0.0000 [-0.0000, 0.0000] | 1.0000 [1.0000, 1.0000] |
| Lasso | -0.0000 [-0.0000, 0.0000] | 1.0000 [1.0000, 1.0000] |
| MLP | 0.0000 [-0.0000, 0.0000] | 1.0000 [1.0000, 1.0000] |
| Brier Score | E<sub>max</sub> | E<sub>avg</sub> |
| 0.2162 [0.2095, 0.2228] | 0.3781 [0.3175, 0.4387] | 0.1622 [0.1500, 0.1743] |
| 0.1999 [0.1939, 0.2059] | 0.3112 [0.2760, 0.3464] | 0.1378 [0.1281, 0.1476] |
| 0.2759 [0.2665, 0.2853] | 0.5464 [0.4694, 0.6233] | 0.2550 [0.2430, 0.2670] |
| Brier Score | E<sub>max</sub> | E<sub>avg</sub> |
| 0.2156 [0.2090, 0.2222] | 0.3526 [0.3088, 0.3963] | 0.1598 [0.1504, 0.1692] |
| 0.1995 [0.1932, 0.2058] | 0.3033 [0.2621, 0.3444] | 0.1355 [0.1231, 0.1478] |
| 0.2742 [0.2634, 0.2850] | 0.5223 [0.4877, 0.5569] | 0.2540 [0.2346, 0.2733] |
| Brier Score | E<sub>max</sub> | E<sub>avg</sub> |
| 0.1887 [0.1848, 0.1926] | 0.2810 [0.1672, 0.3949] | 0.0847 [0.0648, 0.1045] |
| 0.1834 [0.1788, 0.1879] | 0.2456 [0.0911, 0.4002] | 0.0799 [0.0456, 0.1143] |
| 0.1905 [0.1867, 0.1942] | 0.1672 [0.1118, 0.2227] | 0.0565 [0.0400, 0.0729] |

Without any recalibration applied, all models presented varying degrees of miscalibration. The calibration intercepts for each model were negative. LR had an intercept of −0.5298, Lasso −0.4741, and MLP −0.5889, indicating an underestimation of the predicted risks. More importantly, the calibration slopes were significantly below the ideal value of 1.0, suggesting that the models were underfitting the observed data. Among these models, MLP showed the poorest slope at 0.0754 (with a confidence interval of [0.0634, 0.0874]), indicating severe miscalibration. The Brier score, which measures the average squared error of the predicted probabilities, was the highest for MLP at 0.2759, followed by LR at 0.2162 and Lasso at 0.1999. Similarly, Emax, which represents the maximum vertical deviation from perfect calibration, was also highest for MLP at 0.5464, indicating significant local errors in the predicted risk. The Eavg further suggested calibration errors, with MLP again showing the worst performance at 0.2550, while Lasso showed the best performance among the three with an Eavg of 0.1378.

The recalibration in the large adjusts only the intercept while keeping the slope fixed. This approach resulted in modest improvements in the calibration intercepts in all models, but did not affect the slope and moderate calibration metrics. For instance, the calibration intercepts became slightly less negative: LR improved from −0.5298 to −0.5245, and Lasso improved from −0.4741 to −0.4670. As expected, the slopes remained unchanged from the norecalibration setting, with the MLP slope still at 0.0754. Moderate metrics showed slight improvements, with Lasso’s Brier score decreasing to 0.1995 and Eavg improving to 0.1355. Nonetheless, all models still showed poor calibration, particularly MLP, which maintained high scores with a Brier score of 0.2742, Emax of 0.5223, and Eavg of 0.2540.

Logistic recalibration, which includes recalculating the intercept and slope, significantly improved the calibration of all models. All models achieved a perfect weak calibration, with intercepts of 0.0000 and slopes of 1.0000. The moderate calibration metrics also showed significant improvement: Brier scores decreased in all models, with values of 0.1887 for LR, 0.1834 for Lasso, and 0.1905 for MLP. Emax was significantly reduced, especially for MLP, which had a value of 0.1672 (with a confidence interval of [0.1118, 0.2227]). This indicates an improvement in local agreement between predicted and observed probabilities. Eavg, the most informative summary of moderate calibration, showed considerable improvements. MLP outperformed the other models with an Eavg of 0.0565 (confidence interval [0.0400, 0.0729]), followed by Lasso at 0.0799 and LR at 0.0847.

### 3.5. Calibration disparities across gender, race, and ethnicity subgroups

We assessed the calibration of three prediction models, including LR, Lasso, and MLP, across gender, race, and ethnicity subgroups. Without any recalibration applied, LR showed significant miscalibration in several subgroups, as shown in Table 9. For example, the Asian group showed a low calibration slope of 0.3372 and a calibration intercept of −1.0188, indicating severe miscalibration. Similarly, the Hispanic group had a high intercept of 3.0846, with a slope close to 1 but with considerable uncertainty. The other race group showed large deviations, with an intercept of 1.2295 and a slope of 0.5002. No significant differences between the gender groups were evident. For example, the female subgroup had a slope of 0.3173, while the male subgroup had a lower slope of 0.2724. After applying logistic recalibration, all groups achieved perfect calibration, with intercepts close to 0 and slopes close to 1.

**Table 9:** Calibration statistics for LR by gender, race, and ethnicity.

| Model | Group | Method | Calibration Intercept | Calibration Slope |
| --- | --- | --- | --- | --- |
| LR | Male | No Recalibration | -0.4142 [-0.5044, -0.3241] | 0.2724 [0.2233, 0.3215] |
| LR | Male | Recalibration In the Large | -0.4386 [-0.4948, -0.3825] | 0.2724 [0.2233, 0.3215] |
| LR | Male | Logistic Recalibration | -0.0000 [-0.0000, -0.0000] | 1.0000 [1.0000, 1.0000] |
| LR | Female | No Recalibration | -0.5517 [-0.6217, -0.4817] | 0.3173 [0.2873, 0.3473] |
| LR | Female | Recalibration In the Large | -0.5115 [-0.5533, -0.4698] | 0.3173 [0.2873, 0.3473] |
| LR | Female | Logistic Recalibration | 0.0000 [-0.0000, 0.0000] | 1.0000 [1.0000, 1.0000] |
| LR | White | No Recalibration | -0.4673 [-0.5222, -0.4123] | 0.2966 [0.2524, 0.3408] |
| LR | White | Recalibration In the Large | -0.4634 [-0.5115, -0.4153] | 0.2966 [0.2524, 0.3408] |
| LR | White | Logistic Recalibration | -0.0000 [-0.0000, 0.0000] | 1.0000 [1.0000, 1.0000] |
| LR | Black | No Recalibration | -0.6391 [-0.7087, -0.5694] | 0.2821 [0.2249, 0.3394] |
| LR | Black | Recalibration In the Large | -0.5836 [-0.6481, -0.5191] | 0.2821 [0.2249, 0.3394] |
| LR | Black | Logistic Recalibration | 0.0000 [0.0000, 0.0000] | 1.0000 [1.0000, 1.0000] |
| LR | Asian | No Recalibration | -1.0188 [-1.4369, -0.6007] | 0.3372 [0.1215, 0.5528] |
| LR | Asian | Recalibration In the Large | -0.7751 [-1.1026, -0.4476] | 0.3372 [0.1215, 0.5528] |
| LR | Asian | Logistic Recalibration | 0.0000 [-0.0000, 0.0000] | 1.0000 [1.0000, 1.0000] |
| LR | Other | No Recalibration | 1.2295 [0.7193, 1.7397] | 0.5002 [0.2754, 0.7249] |
| LR | Other | Recalibration In the Large | 0.2120 [0.0888, 0.3351] | 0.5002 [0.2754, 0.7249] |
| LR | Other | Logistic Recalibration | 0.0000 [-0.0000, 0.0000] | 1.0000 [1.0000, 1.0000] |
| LR | Hispanic | No Recalibration | 3.0846 [1.2873, 4.8819] | 0.9949 [0.0448, 1.9450] |
| LR | Hispanic | Recalibration In the Large | 0.4358 [-0.5756, 1.4471] | 0.9949 [0.0448, 1.9450] |
| LR | Hispanic | Logistic Recalibration | 0.0000 [-0.0000, 0.0000] | 1.0000 [1.0000, 1.0000] |
| LR | Non-Hispanic | No Recalibration | -0.5712 [-0.6143, -0.5281] | 0.2836 [0.2450, 0.3223] |
| LR | Non-Hispanic | Recalibration In the Large | -0.5355 [-0.5759, -0.4951] | 0.2836 [0.2450, 0.3223] |
| LR | Non-Hispanic | Logistic Recalibration | 0.0000 [-0.0000, 0.0000] | 1.0000 [1.0000, 1.0000] |
| LR | Other | No Recalibration | 0.1191 [-0.2316, 0.4698] | 0.5062 [0.3477, 0.6646] |
| LR | Other | Recalibration In the Large | -0.1889 [-0.2768, -0.1011] | 0.5062 [0.3477, 0.6646] |
| LR | Other | Logistic Recalibration | -0.0000 [-0.0000, 0.0000] | 1.0000 [1.0000, 1.0000] |

Similar to LR, Lasso presented similar results, with the Asian, Hispanic, and other race groups again showing the most significant miscalibration, as shown in Table 10. The Asian group had a calibration intercept of −0.9354 and a slope of 0.4823, while the Hispanic group had a high intercept of 3.0619 and a slope exceeding 1. The other race group also showed poor calibration, with an intercept of 1.3131 and a slope of 0.6494. In contrast, the calibration performance among the gender and white and non-Hispanic subgroups was more stable but still suboptimal. Logistic recalibration successfully corrected these biases, yielding intercepts and slopes approaching ideal values across all groups.

**Table 10:** Calibration statistics for Lasso by gender, race, and ethnicity.

| Model | Group | Method | Calibration Intercept | Calibration Slope |
| --- | --- | --- | --- | --- |
| Lasso | Male | No Recalibration | -0.3420 [-0.4418, -0.2423] | 0.3686 [0.3051, 0.4321] |
| Lasso | Male | Recalibration In the Large | -0.3805 [-0.4393, -0.3218] | 0.3686 [0.3051, 0.4321] |
| Lasso | Male | Logistic Recalibration | 0.0000 [-0.0000, 0.0000] | 1.0000 [1.0000, 1.0000] |
| Lasso | Female | No Recalibration | -0.4818 [-0.5583, -0.4053] | 0.4264 [0.3821, 0.4707] |
| Lasso | Female | Recalibration In the Large | -0.4297 [-0.4806, -0.3788] | 0.4264 [0.3821, 0.4707] |
| Lasso | Female | Logistic Recalibration | -0.0000 [-0.0000, 0.0000] | 1.0000 [1.0000, 1.0000] |
| Lasso | White | No Recalibration | -0.4005 [-0.4644, -0.3365] | 0.3969 [0.3350, 0.4587] |
| Lasso | White | Recalibration In the Large | -0.3990 [-0.4500, -0.3479] | 0.3969 [0.3350, 0.4587] |
| Lasso | White | Logistic Recalibration | -0.0000 [-0.0000, 0.0000] | 1.0000 [1.0000, 1.0000] |
| Lasso | Black | No Recalibration | -0.5578 [-0.6429, -0.4727] | 0.3899 [0.3065, 0.4733] |
| Lasso | Black | Recalibration In the Large | -0.4884 [-0.5665, -0.4104] | 0.3899 [0.3065, 0.4733] |
| Lasso | Black | Logistic Recalibration | -0.0000 [-0.0000, 0.0000] | 1.0000 [1.0000, 1.0000] |
| Lasso | Asian | No Recalibration | -0.9354 [-1.3397, -0.5312] | 0.4823 [0.2600, 0.7045] |
| Lasso | Asian | Recalibration In the Large | -0.6112 [-0.9100, -0.3125] | 0.4823 [0.2600, 0.7045] |
| Lasso | Asian | Logistic Recalibration | 0.0000 [-0.0000, 0.0000] | 1.0000 [1.0000, 1.0000] |
| Lasso | Other | No Recalibration | 1.3131 [0.7690, 1.8572] | 0.6494 [0.3640, 0.9347] |
| Lasso | Other | Recalibration In the Large | 0.1391 [0.0040, 0.2742] | 0.6494 [0.3640, 0.9347] |
| Lasso | Other | Logistic Recalibration | -0.0000 [-0.0000, 0.0000] | 1.0000 [1.0000, 1.0000] |
| Lasso | Hispanic | No Recalibration | 3.0619 [1.3422, 4.7816] | 1.0988 [0.0643, 2.1334] |
| Lasso | Hispanic | Recalibration In the Large | 0.4017 [-0.7019, 1.5054] | 1.0988 [0.0643, 2.1334] |
| Lasso | Hispanic | Logistic Recalibration | -0.0000 [-0.0000, 0.0000] | 1.0000 [1.0000, 1.0000] |
| Lasso | Non-Hispanic | No Recalibration | -0.5003 [-0.5508, -0.4498] | 0.3865 [0.3338, 0.4392] |
| Lasso | Non-Hispanic | Recalibration In the Large | -0.4576 [-0.5064, -0.4089] | 0.3865 [0.3338, 0.4392] |
| Lasso | Non-Hispanic | Logistic Recalibration | -0.0000 [-0.0000, 0.0000] | 1.0000 [1.0000, 1.0000] |
| Lasso | Other | No Recalibration | 0.2198 [-0.2338, 0.6734] | 0.6599 [0.4007, 0.9192] |
| Lasso | Other | Recalibration In the Large | -0.1408 [-0.2408, -0.0408] | 0.6599 [0.4007, 0.9192] |
| Lasso | Other | Logistic Recalibration | 0.0000 [-0.0000, 0.0000] | 1.0000 [1.0000, 1.0000] |

Among the three models, MLP had the poorest calibration performance prior to recalibration, as shown in Table 11. The calibration slopes were consistently low across all subgroups, typically around 0.07 (for example, 0.0731 for the male group and 0.0629 for the Asian group). The calibration intercepts were also extreme scores, with an intercept of −1.0672 for the Asian group and an intercept of 1.9491 for the Hispanic group, reflecting underconfidence and overconfidence in predicted probabilities. Logistic recalibration again mitigated these issues, improving intercepts and slopes across all subgroups.

**Table 11:** Calibration statistics for MLP by gender, race, and ethnicity.

| Model | Group | Method | Calibration Intercept | Calibration Slope |
| --- | --- | --- | --- | --- |
| MLP | Male | No Recalibration | -0.4469 [-0.5183, -0.3756] | 0.0731 [0.0642, 0.0820] |
| MLP | Male | Recalibration In the Large | -0.5174 [-0.5659, -0.4690] | 0.0731 [0.0642, 0.0820] |
| MLP | Male | Logistic Recalibration | 0.0000 [-0.0000, 0.0000] | 1.0000 [1.0000, 1.0000] |
| MLP | Female | No Recalibration | -0.6373 [-0.7155, -0.5592] | 0.0676 [0.0556, 0.0795] |
| MLP | Female | Recalibration In the Large | -0.6672 [-0.7192, -0.6151] | 0.0676 [0.0556, 0.0795] |
| MLP | Female | Logistic Recalibration | -0.0000 [-0.0000, 0.0000] | 1.0000 [1.0000, 1.0000] |
| MLP | White | No Recalibration | -0.5273 [-0.5848, -0.4697] | 0.0698 [0.0623, 0.0772] |
| MLP | White | Recalibration In the Large | -0.5796 [-0.6245, -0.5348] | 0.0698 [0.0623, 0.0772] |
| MLP | White | Logistic Recalibration | 0.0000 [-0.0000, 0.0000] | 1.0000 [1.0000, 1.0000] |
| MLP | Black | No Recalibration | -0.6795 [-0.7726, -0.5863] | 0.0710 [0.0582, 0.0837] |
| MLP | Black | Recalibration In the Large | -0.7008 [-0.7586, -0.6431] | 0.0710 [0.0582, 0.0837] |
| MLP | Black | Logistic Recalibration | -0.0000 [-0.0000, 0.0000] | 1.0000 [1.0000, 1.0000] |
| MLP | Asian | No Recalibration | -1.0672 [-1.4289, -0.7055] | 0.0629 [0.0279, 0.0979] |
| MLP | Asian | Recalibration In the Large | -0.9765 [-1.2430, -0.7101] | 0.0629 [0.0279, 0.0979] |
| MLP | Asian | Logistic Recalibration | -0.0000 [-0.0000, 0.0000] | 1.0000 [1.0000, 1.0000] |
| MLP | Other | No Recalibration | 1.0316 [0.4635, 1.5997] | 0.1106 [0.0494, 0.1717] |
| MLP | Other | Recalibration In the Large | 0.3724 [0.2214, 0.5234] | 0.1106 [0.0494, 0.1717] |
| MLP | Other | Logistic Recalibration | 0.0000 [-0.0000, 0.0000] | 1.0000 [1.0000, 1.0000] |
| MLP | Hispanic | No Recalibration | 1.9491 [1.4201, 2.4781] | 0.1121 [0.0506, 0.1736] |
| MLP | Hispanic | Recalibration In the Large | 0.9655 [0.4468, 1.4843] | 0.1121 [0.0506, 0.1736] |
| MLP | Hispanic | Logistic Recalibration | 0.0000 [0.0000, 0.0000] | 1.0000 [1.0000, 1.0000] |
| MLP | Non-Hispanic | No Recalibration | -0.6256 [-0.6682, -0.5829] | 0.0689 [0.0622, 0.0756] |
| MLP | Non-Hispanic | Recalibration In the Large | -0.6535 [-0.6745, -0.6325] | 0.0689 [0.0622, 0.0756] |
| MLP | Non-Hispanic | Logistic Recalibration | 0.0000 [-0.0000, 0.0000] | 1.0000 [1.0000, 1.0000] |
| MLP | Other | No Recalibration | -0.0867 [-0.3133, 0.1398] | 0.0855 [0.0689, 0.1021] |
| MLP | Other | Recalibration In the Large | -0.2906 [-0.3859, -0.1952] | 0.0855 [0.0689, 0.1021] |
| MLP | Other | Logistic Recalibration | -0.0000 [-0.0000, 0.0000] | 1.0000 [1.0000, 1.0000] |

## 4. Discussion

In this study, we used a large cohort of 4,503,509 patients from the One-Florida dataset to develop and validate a risk prediction tool for the early detection of HF onset. From this cohort, we identified 2,561 HF cases and 5,493 matched controls. We employed a suite of NLP models that generated dense vector representations of clinical concepts, which were used as inputs for five machine learning models. The results of this study indicate that the XGBoost model outperformed others in identifying patients at high risk for HF onset. We performed a comprehensive feature importance analysis, which suggested that diagnoses, procedures, medications, lab tests, and patient age were among the most influential predictors. These findings provide valuable insights into underrecognized or emerging risk factors and have the potential to inform more targeted HF screening strategies.

We further assessed the performance of the model across subgroups defined by gender, race, and ethnicity, revealing significant variations in the prediction results. Within linear models, Lasso outperformed LR, offering better discrimination, calibration, and overall predictive accuracy, especially within the Hispanic subgroup. This finding suggests that Lasso’s built-in regularization is more likely to capture key predictors, especially in contexts with high-dimensional and sparse EHR data. Despite Lasso’s improved prediction performance, the observed disparities across subgroups highlight the need for further investigation into model robustness and generalizability, especially for underrepresented groups.

In tree-based models, XGBoost outperformed RF, achieving the highest performance in the Hispanic subgroup. The superior performance of XGBoost can be attributed to its ability to model complex nonlinear interactions. Notably, the relatively lower performance for the Black subgroup, mainly when using RF, raises concerns about potential biases in the training data. This result highlights the need for targeted efforts to ensure equitable model training, such as subgroup-specific data augmentation and reweighting techniques.

Among the neural network models, MLP consistently outperformed GRU and RETAIN across all subgroups and evaluation metrics, indicating its effectiveness and robustness. While GRU and RETAIN presented competitive AUROC and precision in certain subgroups, their performance was generally less consistent, potentially due to suboptimal sequence modeling in high-dimensional and sparse EHR data. In contrast, MLP achieved a more balanced trade-off between precision and recall, resulting in higher F1 scores and better calibration. A key factor contributing to the strong performance of MLP is its use of dense vector representations derived from structured medical codes. These embeddings transform sparse, high-dimensional categorical inputs (i.e., structured medical codes) into continuous, low-dimensional representations that capture the underlying semantic relationships between clinical concepts. By utilizing these informative feature spaces, the MLP can mitigate the impacts of data sparsity and noise, improving its predictive accuracy.

The calibration results revealed significant differences in how each model estimates risk and responds to recalibration strategies. Without recalibration, all models (LR, Lasso, and MLP) displayed varying levels of miscalibration, with MLP performing the poorest performance in both weak and moderate calibration. The observed negative intercepts across the models suggest a systematic underestimation of risk, while slopes significantly below 1, especially in the case of MLP, indicate underfitting and inadequate discrimination. These findings highlight the difficulties associated with achieving well-calibrated probability estimations from neural network models without incorporating additional calibration strategies. There are similarities between the attitudes expressed by the importance of posthoc recalibration in this study and those described by previous research [27–29].

Recalibration in the large, which adjusts only the average prediction bias (i.e., the intercept), led to marginal improvements and failed to address the issue of miscalibrated spread (i.e., slope). In contrast, logistic recalibration led to significant improvements in both weak and moderate calibration for all models, effectively aligning predicted probabilities with observed outcomes. Notably, MLP, which initially showed the poorest calibration, benefited the most from logistic recalibration and achieved the lowest Eavg, a key indicator of overall calibration quality. Moreover, these improvements were consistently observed across gender, race, and ethnicity subgroups, reducing disparities and aligning predicted probabilities more closely with observed outcomes.

## 5. Study limitations

Several limitations to this study need to be acknowledged. First, EHR data used for model development and evaluation were collected from a single institution. This may limit the applicability of our model to broader populations of HF and diverse clinical settings with varying demographics and care protocols. Second, EHR data are inherently noisy and often contain missing, incomplete, or inaccurately coded information. These issues are often due to variations in documentation practices, coding errors, and inconsistencies in data capture. Such factors could introduce biases that affect the reliability of our method. Lastly, the lack of an external validation cohort adds further caution regarding the generalizability of these findings. While internal validation offers insight into the predictive power of the model, external validation across multiple institutions with heterogeneous populations is critically important for assessing the adaptability of the model in real-world clinical scenarios.

## 6. Conclusions

Early detection of HF onset is crucial for enabling timely and personalized preventive interventions that can improve patient outcomes and reduce healthcare costs. Emerging advances in deep learning techniques, particularly NLP-based approaches, have offered new opportunities for facilitating the accurate targeting of patients at high risk of developing HF. In this study, we introduced an NLP-based approach for risk stratification, utilizing a large amount of EHR data from a single medical institution. Our approach demonstrated strong predictive accuracy and clinical relevance, highlighting the potential of integrating NLP-driven risk stratification tools into realworld clinical practices. Further studies need to be carried out in order to validate the effectiveness of this approach across multiple healthcare systems and patient populations.

## Data Availability

All data produced in the present work are contained in the manuscript

## Declaration of Competing Interest

The authors declare no conflict of interests.

## Notes

### Competing Interest Statement

The authors have declared no competing interest.

### Funding Statement

This study was funded by NIH grants R01AG083039, RF1AG084178, RF1AG077820, R01AG080991, R01AG080624, and R01AG076234.

### Author Declarations

Ethics committee/IRB of the University of Florida gave ethical approval for this work.

